# Detection of Epileptic Spasms Using Foundational AI and Smartphone Videos: A Novel Diagnostic Approach for a Rare Neurological Disorder

**DOI:** 10.1101/2024.10.28.24316130

**Authors:** Gadi Miron, Mustafa Halimeh, Simon Tietze, Martin Holtkamp, Christian Meisel

## Abstract

Infantile epileptic spasm syndrome (IESS) is a severe neurological disorder characterized by epileptic spasms (ES). Timely diagnosis and treatment are crucial but often delayed due to symptom misidentification. Smartphone videos can aid in diagnosis, but availability of specialist review is limited. We fine-tuned a foundational video model for ES detection using social media videos, thus addressing this clinical need and the challenge of data scarcity in rare disorders. Our model, trained on 141 children with 991 seizures and 127 children without seizures, achieved high performance (area under the receiver-operating-curve (AUC) 0.96, 83% sensitivity, 95% specificity) including validation on external datasets from smartphone videos (93 children, 70 seizures, AUC 0.98, false alarm rate (FAR) 0.75%) and gold-standard video-EEG (22 children, 45 seizures, AUC 0.98, FAR 3.4%). This study demonstrates the potential of smartphone videos for AI-powered analysis as the basis for accelerated IESS diagnosis and novel strategy for diagnosis of rare disorders.

## Introduction

Timely diagnosis of rare neurological disorders remains a significant challenge in healthcare, often resulting in delayed patient treatment^1^. Infantile epileptic spasm syndrome (IESS), a developmental and epileptic encephalopathy affecting approximately 1 in 2000-2500 infants in their first year of life, exemplifies this problem^2,3^. While approximately 9% of children experience some sort of paroxysmal movement events during their first year of life (most of which are benign), a smaller percentage actually have seizures^4^. Despite the stereotypical nature of the hallmark seizures of IESS, epileptic spasms (ES), diagnosis is frequently delayed by weeks to months due to misidentification of symptoms as benign physiological occurrences or failure to recognize any abnormality by physicians or parents ^5–12^. These delays are associated with long-term poor cognitive outcomes, inadequate seizure control, increased disability, and higher healthcare costs^7,13^.

The widespread availability of smartphones and advancements in artificial intelligence (AI) may open new avenues for digital health technologies and accelerated neurological diagnostics. In epilepsy, videos captured by smartphones have already been shown to enhance diagnostic accuracy and clinical decision-making while reducing patient and family stress^14^. In the hands of experts, these videos facilitate earlier arrival to clinics, faster diagnostic EEGs, and improved treatment responses for children with IESS^15^. Videos from smartphones have furthermore been shown to be at times non-inferior to gold standard video-EEG monitoring for initial diagnosis, offering advantages particularly in resource-limited settings^16,17^. Thus, while smartphone videos could in principle enable more rapid and accurate identification of ES, the shortage of medical professionals available for timely review and evaluation of patient videos limits the broader applicability of this approach. Rapid video evaluations supported by AI may help to broadly scale expert knowledge in detecting paroxysmal movements suspicious for seizures in infants and young children as basis for faster diagnosis and treatment. Developing accurate AI models for rare conditions like IESS, however, presents unique challenges, primarily due to the scarcity of large, labeled datasets required for effective model training.

Here, we address the challenge of timely IESS diagnosis with smartphone-based detection of ES by leveraging two key developments: powerful foundation vision models pretrained on extensive datasets and utilization of the wealth of publicly available video data in social media. First, foundation vision models based on transformer architectures, trained on extensive datasets of images and videos from the internet, have been robust for human activity recognition^18,19^. These models have potential to significantly enhance video-based seizure detection, extending capabilities to additional video sources including smartphone recordings. This is particularly valuable given the variability in video quality, recording equipment, and patient demographics encountered in real-world settings. Second, social media platforms have inadvertently become valuable repositories of medical information, with user-uploaded videos clearly demonstrating the semiology of stereotypical seizure disorders. While medical research has begun to leverage social media data for various purposes, this approach remains largely unexplored for rare neurological disorders and epilepsy. This untapped resource offers a potential solution to the data scarcity problem, enabling the development of robust AI models for conditions that traditionally lack sufficient clinical data. We here curated a large dataset of ES seizures from open-source videos, thus addressing the data scarcity issue. We then trained an AI model for automated detection of ES, building on a state-of-the-art vision foundational model, thus benefitting from comprehensive pretraining. Finally, we validated our model on three additional independent datasets, comprising both smartphone recordings and gold standard in-hospital video-EEG monitoring data, thus ensuring model robustness across varied video sources and clinical settings.

## Methods

### Derivation dataset collection and annotation

The study design, data collection, preprocessing, AI model development and testing are depicted in Figure 1. We conducted a systematic search on YouTube videos published before 2022 using the key words “infantile spasms”, “epileptic spasms”, and “west syndrome”. Videos were included based on the following criteria: (1) subject appears to be under 2 years of age, (2) video contains an event consistent with ES semiology, as independently confirmed by two expert neurologists (GM and CM), (3) subject is clearly visible and not substantially obstructed by other people, objects or overlying text, (4) video quality (resolution, lighting) is sufficient for visual recognition of semiology, without obscuring filters or effects. Exclusion criteria were: (1) subject or limbs obstructed from view (e.g., close ups of the face or trunk), (2) insufficient video quality for semiology determination, (3) subject is clearly older than 2 years. To ensure a diverse population, videos were included regardless of the country of origin, language spoken, video length, or upload year. Next, 5-second segments were annotated, either containing stereotypical movements for ES or non-seizure segments.

**Figure 1.**
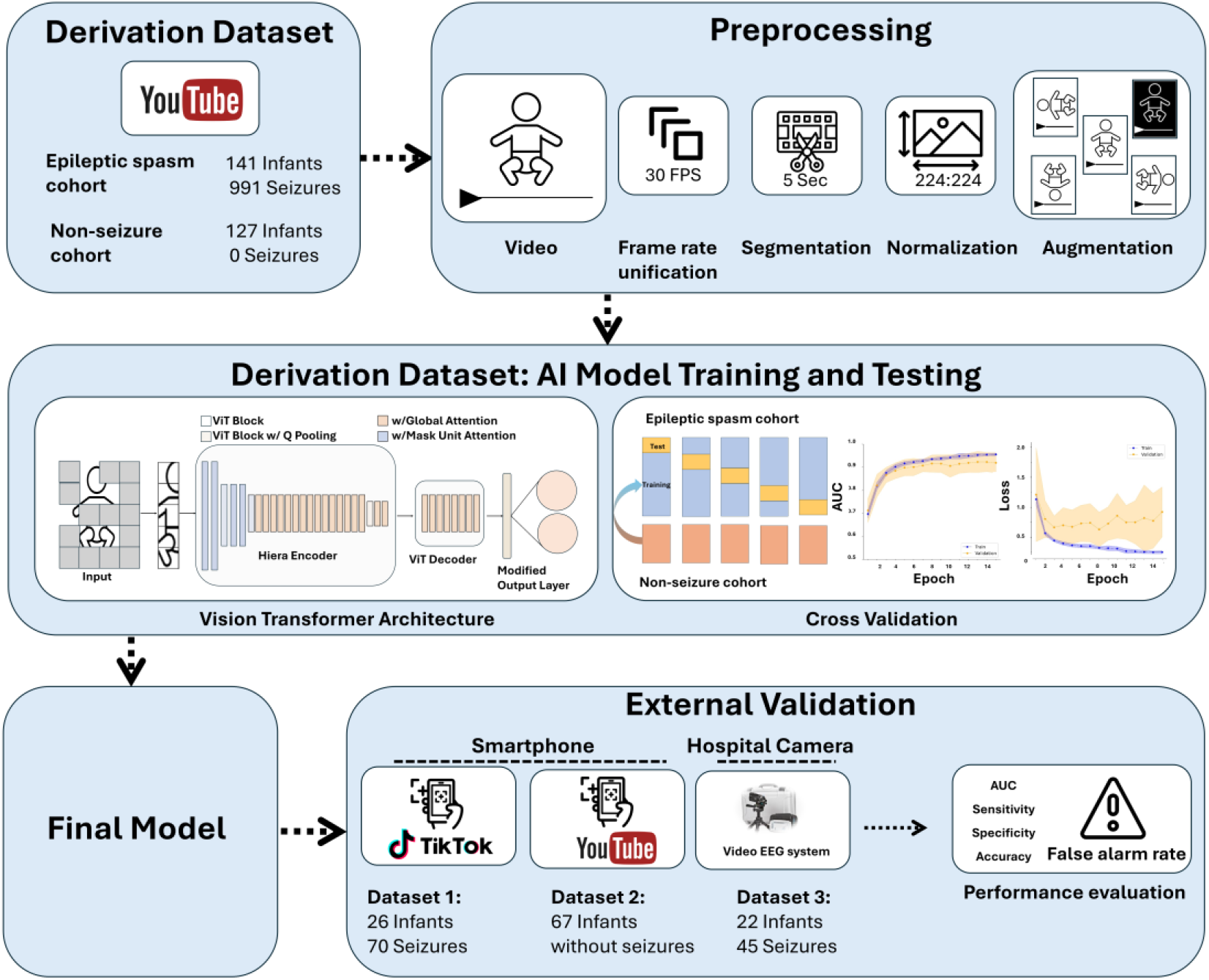
Study design. To acquire the derivation dataset, we conducted a search for YouTube videos with the key words “epileptic spasms”, “infantile spasms”, and “West syndrome”. All video segments were reviewed by expert neurologists and underwent preprocessing. Videos were analyzed by a vision transformer model that was re-trained for the purpose of epileptic spasm identification. A five-fold cross validation approach was used for training and testing. Finally, one single model was derived from the entire derivation dataset and tested on three external validation datasets: two datasets sourced from smartphone videos, and one dataset from gold-standard video-EEG monitoring. Performance on these validation datasets was assessed using the area under the receiver-operating-curve (AUC), sensitivity, specificity, accuracy and false alarm rate.

To further enhance our training dataset, we incorporated additional videos of normally behaving infants from previously collected YouTube datasets(24–26). All segments from these datasets were manually reviewed to confirm the absence of seizures while adhering to our inclusion criteria.

### Out-of-sample external testing

We collected three additional external datasets to further validate seizure detection performance and evaluate false alarm rate (FAR): (1) smartphone videos of infants with ES published on YouTube after 2022 and on TikTok, collected using the same approach as for the derivation dataset. (2) An additional cohort of normally behaving infants from YouTube to assess FAR on smartphones. (3) Videos from long-term video-EEG monitoring recordings of infants under two years age at the Epilepsy-Center Berlin-Brandenburg. For each video, we included all 5-seconds segments where the subject was clearly visible. Since in-hospital videos from long-term video-EEG monitoring were of long duration and captured with stationary cameras with a wide view, these videos were cropped automatically around the infant using an existing AI model designed for object detection^20^. For all in-hospital video segments used for FAR evaluation, seizure activity was ruled out by video-EEG.

### Meta-information and technical characteristics analysis

When available, we collected additional meta-information from the online videos of ES, including view count, likes, comments, and upload year, to assess the diversity and reach of this content. To evaluate the technical heterogeneity of the collected videos, we analyzed video resolution, duration, frame rate, and bitrate using python library OpenCV and FFmpeg. Additionally, we quantified general visual characteristics by analyzing brightness, daytime vs. nighttime recordings, motion, and sharpness of videos. Brightness was calculated as the mean pixel intensity across sampled frames, providing insight into lighting conditions. Nighttime recording was estimated using the averaged variance of the RGB pixels, if the average was below a predefined threshold, then the video was classified as recorded during night, otherwise during day. Motion was assessed by measuring the mean absolute difference between consecutive frames, indicating the level of movement or camera stability. Sharpness was evaluated using the variance of the Laplacian operator, offering a measure of image clarity.

### Video data preprocessing

To ensure consistency and to enhance the dataset for model training, we performed the following preprocessing steps: (1) we unified frame rates to 30 frames per second using FFmpeg. (2) We cut videos into 5-second segments, annotating each as seizure or non-seizure. (3) We applied several augmentations to each video segment, including vertical flip, horizontal flip, 90-degree clockwise and counterclockwise rotation, and color inversion to increase variability within our dataset. (4) We standardized frame size to 224 by 224 pixels by downsampling the resolution of each frame using a trilinear interpolation function. (5) We normalized color values per pixel using z-transformation to meet the requirements of the foundational model used^19^.

### Training and testing of classification model

We trained our model using the Hiera Vision Transformer, which was pretrained on the Kinetics 400 Human Action Recognition dataset^18,19^. The model was adapted for our purpose by modifying the final classification layer for binary classification using sigmoid function, distinguishing between seizure and non-seizure video segments. We used a parameter fine-tuning method to then train the model for seizure detection. We employed a 5-fold cross-validation approach for training and testing on the derivation dataset. The data was split at the child level into 5 non-overlapping test folds, ensuring all segments from a single child were in the same fold. We trained the model for 15 epochs with a batch size of 4, using a learning rate that started at 0.0001 and was multiplied by 0.75 every 10 epochs. To improve specificity, additional data from 127 healthy infants were included in the training set only. Finally, we performed additional evaluations of model performance on three independent datasets, using a classification model trained on the entire derivation dataset.

### Performance metrics

We assessed the model’s performance using area under the receiver-operating-characteristic curve (AUC), sensitivity, specificity, accuracy, and false alarm rate (FAR). For children who had only one class of video segments (i.e., either only seizure segments or only non-seizure video segments), AUC and the metric measuring the missed segment class were not calculated. We used the standard threshold of 0.5 to calculate sensitivity, specificity, and accuracy. We reported the FAR, evaluated on seizure-free children, as the percentage of normal segments incorrectly classified as seizures. We present all results using the mean and 95% confidence interval.

## Data availability

Open-source smartphone video URLs are available upon reasonable request. Video-EEG monitoring data is not publicly available due to patient privacy concerns.

## Code availability

The underlying code is not publicly available for proprietary reasons. We used Python 3.9.13 with specific packages (torch 2.2.0, hiera-transformer 0.1.2) on an Intel(R) Xeon(R) Silver 4216 CPU with two NVIDIA RTX A6000 GPUs.

### Ethics Statement

This study was approved by the Institutional Review Board of Charité – Universitätsmedizin Berlin (reference number EA2/273/23).

## Results

### Derivation dataset: population characteristics, training, performance

For model training and testing, we collected videos from 141 children with ES, comprising 991 seizures and 597 non-seizure video segments of 5-second duration each. The median number of segments per child was 6 (IQR 4-12, range 1-42). We further enriched the training dataset with 127 healthy infants, contributing 1385 video segments (median 8 per child, IQR 4-14, range 1-70; Table 1). Videos containing ES were published between 2007 and 2022, with a median of 8,885 views per video (IQR 1,687-42,274.5), 31 likes (IQR 8-121.5) and 5 comments (IQR 1-13.5). As videos were derived from social media, they exhibited considerable technical heterogeneity in terms of resolution, bitrate, brightness of the video, and sharpness (Table 1).

**Table 1.** Study Population and Video Characteristics.

|  | Derivation Dataset |  | External Validation Dataset |  |  |
| --- | --- | --- | --- | --- | --- |
|  | Epileptic Spasm Cohort | Normally Behaving Infants | Dataset 1 - Smartphone - Epileptic Spasm Cohort | Dataset 2 - Smartphone - Normally Behaving Infants | Dataset 3 – Hospital-Video-EEG Monitoring |
| Number of subjects | 141 | 127 | 26 | 67 | 22 |
| Data source | YouTube | YouTube | TikTok, YouTube | YouTube | Video-EEG monitoring |
| Seizure segments | 991 | 0 | 70 | 0 | 45 |
| Non-Seizure segments | 597 | 1385 | 31 | 666 | 11190 |
| Segments per child, median (IQR) | 6 (4-12) | 8 (4-14) | 3 (2-5) | 6 (2.75 - 10.5) | 382 (158.25 - 766) |
| Resolution: high / medium /low% | 57/7/36 % | 52/11/37% | 88/8/4% | 57/34/9% | 2/77/21% |
| Bitrate, median (IQR) kbps | 530 (301–1016) | 949 (449-1637) | 1113 (925-1403) | 2146 (779 - 3243) | 3538 (3404-3671) |
| Sharpness values, median (IQR) laplacian variance | 88 (45–216) | 44 (22-108) | 303 (149-559) | 60 (24-82) | 284 (196-411) |
| Brightness, median (IQR) gray level intensity | 104 (80-123) | 112 (89-138) | 117 (111-133) | 124 (101-143) | 107 (89-125) |

High resolution is defined as above 720p, medium resolution between 480-720p, and low resolution under 480p. Abbreviations: kbps - kilobytes per second; IQR - interquartile (25<sup>th</sup> to 75<sup>th</sup>) range.

Our model detected ES with an AUC of 0.96 (CI 0.94-0.98, five-fold cross-validation) on the derivation dataset. Using a threshold of 0.5, this provided a sensitivity of 82% (CI 78% - 87%), specificity of 90% (CI 86% - 94%), and accuracy of 85% (CI 82%-88%) on the test folds (Figure 2). No statistically significant relationships were found between the technical characteristics of videos and prediction performance in the derivation dataset.

**Figure 2.**
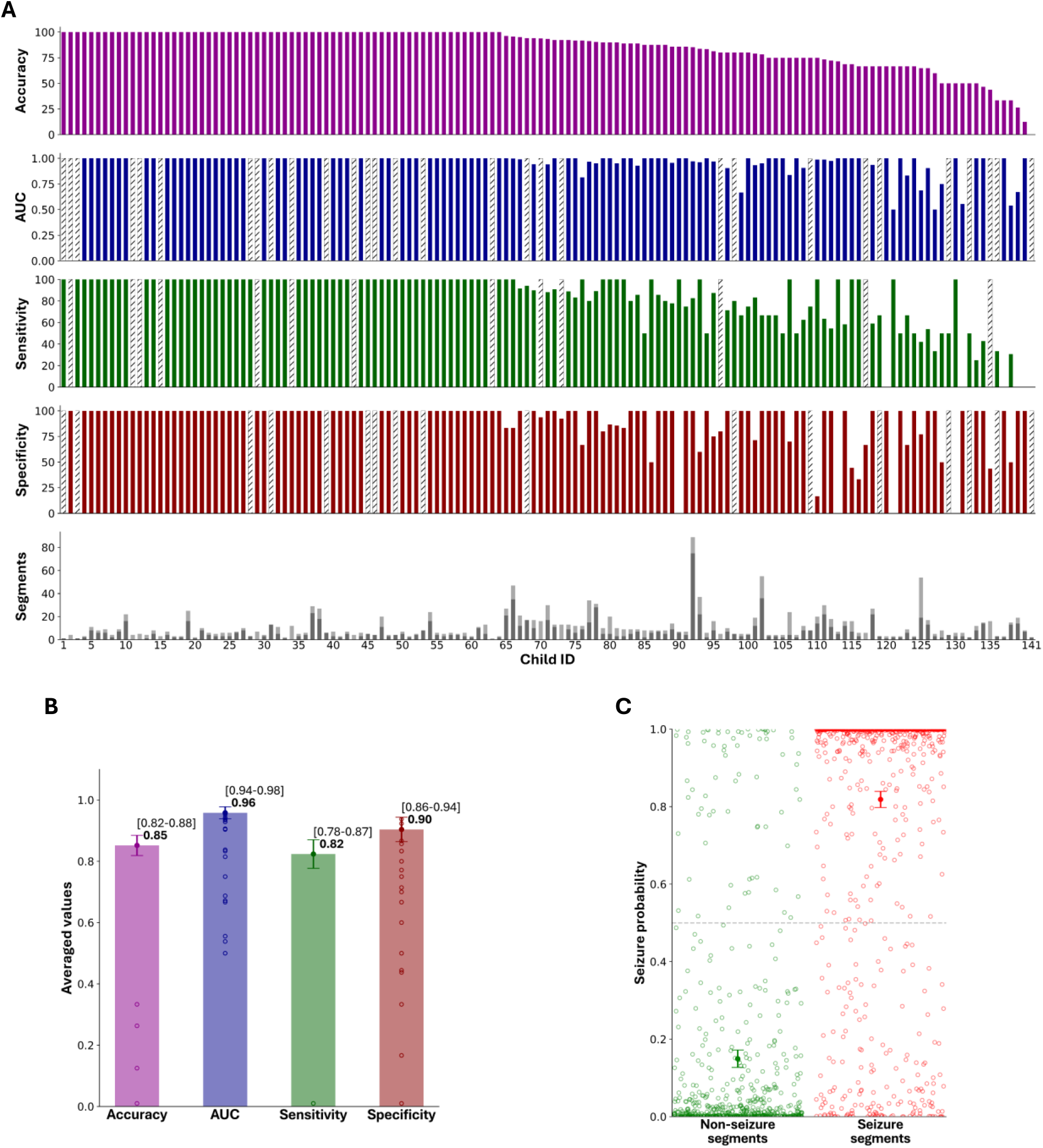
Seizure detection performance on the derivation dataset (smartphone videos). A, Each column represents one subject. Only children that had both seizure and normal segments (111/141) were included in calculating AUC (in cases where only one class of videos was available, a gray column is depicted). B, Overall metrics of performance across all subjects. C, All model predictions, a threshold of 0.5 is depicted as dashed line. Markers denote mean and 95% confidence intervals. Abbreviations: AUC - area under the receiver-operating-characteristic curve.

### External validation: population characteristics, performance, false alarm rate

Using the fixed, trained algorithm with fixed threshold, we next evaluated our seizure detection model on three independent datasets to assess out-of-sample performance, establish FAR, and evaluate transferability to different video sources. Dataset 1, smartphone-based, included 26 infants with ES (70 seizure and 31 non-seizure 5-second video segments). Our model achieved high performance, comparable to the derivation dataset testing, with an AUC of 0.98 (95% CI: 0.94-1.0), sensitivity 89% (95% CI: 82-95%), specificity 100% (95% CI: 100-100%), and accuracy of 92% (95% CI: 87-97%; Figure 3).

**Figure 3.**
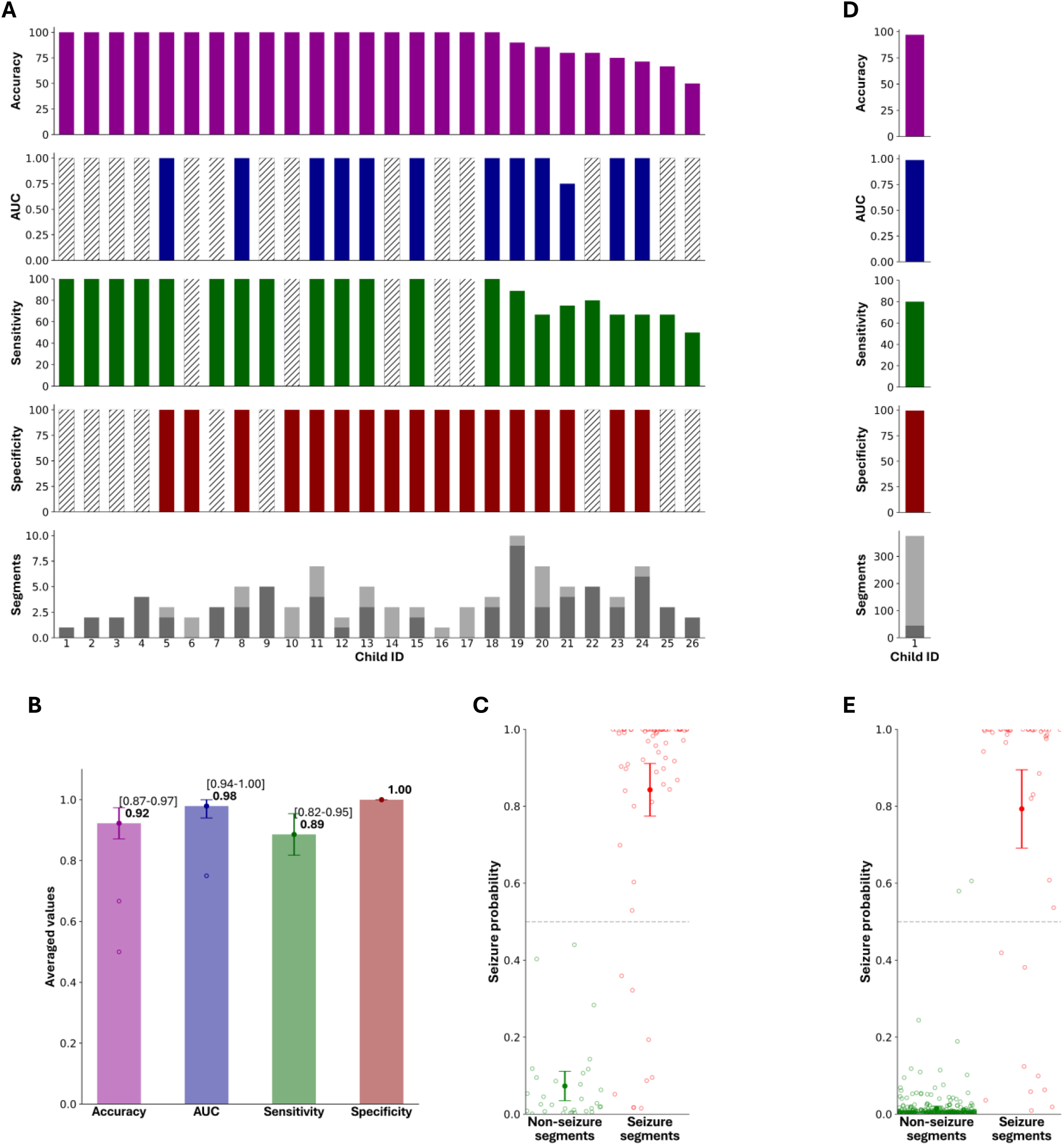
Performance of classification model on external validation datasets (smartphone and gold-standard video-EEG). A, Individual performance metrics for 26 children with epileptic spasms included in the external validation dataset 1 (smartphone-derived), and 1 child with epileptic spasms in external validation dataset 3 (hospital-derived). Each column represents one subject. Only children that had both seizure and normal segments were included in calculating AUC (in cases where only one class of videos was available, a gray column is depicted). B, Overall metrics of performance across all subjects. C, All model predictions are shown. Markers denote mean and 95% confidence intervals. Abbreviations: AUC - area under the receiver-operating-characteristic curve.

Dataset 2, also smartphone-based, included 67 normally behaving infants with 666 5-seconds segments. False detections were identified in 0.75% (5/666) of evaluated video segments, with 62 of 67 subjects having no false alarms. This resulted in a mean FAR per patient of 1.6% (95% CI: 0.0%-3.4%; Figure 4).

**Figure 4.**
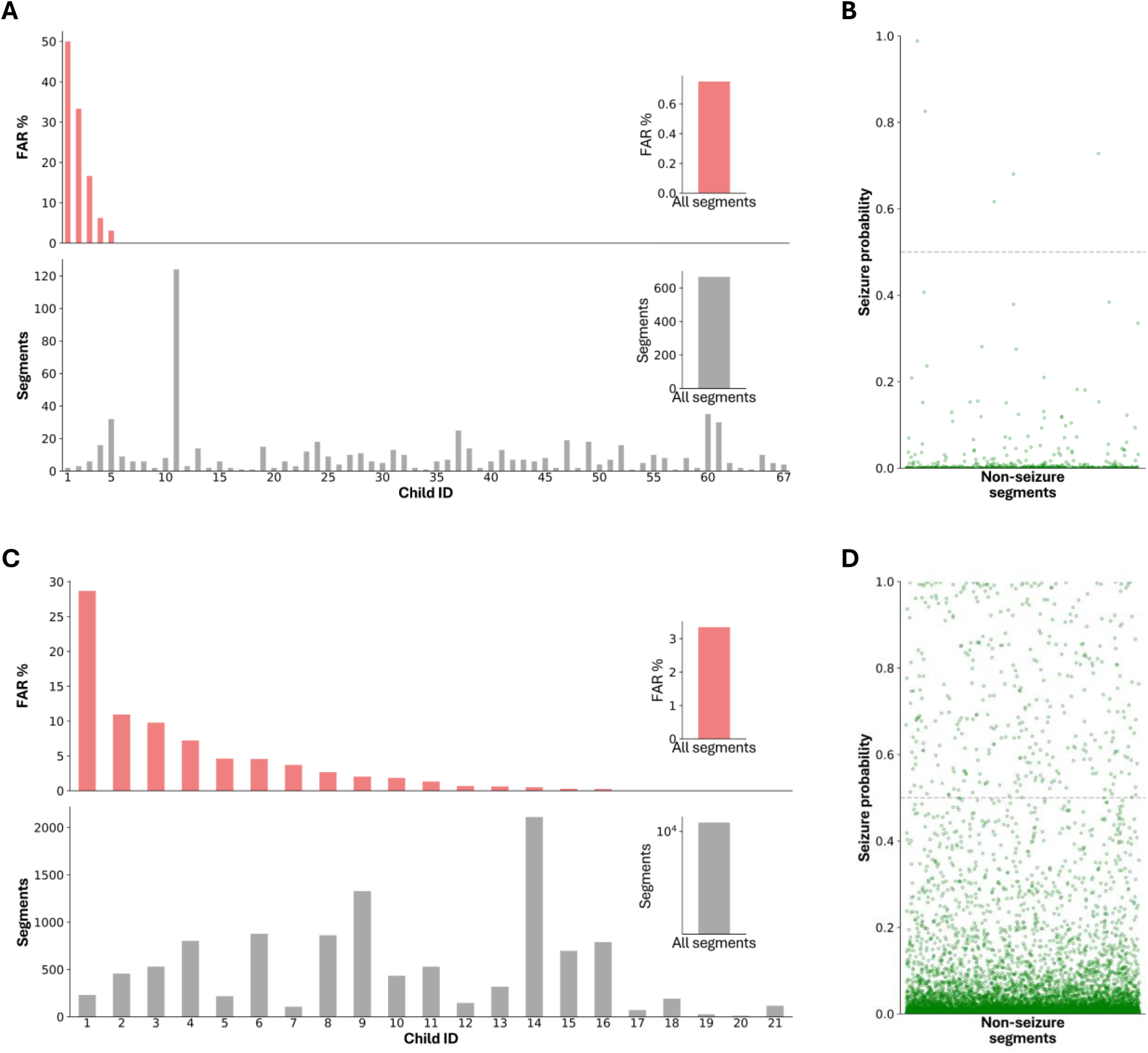
False alarm rate (FAR) evaluation. All subjects evaluated had no seizures, every positive prediction was considered as a false alarm. The FAR is the percentage of positive predictions across all subjects. A, FAR evaluation on an out-of-sample smartphone-derived video dataset including 67 children and 666 five-second segments (dataset 2). Each bar corresponds to one child. In total, 5 of 666 segments were classified as positive detections (0.75%), across all children the average is 1.6% B, All the video segments are plotted with the prediction probability. C, FAR evaluation on an out-of-sample gold standard video EEG derived dataset including 21 children and 10,860 five-second video segments (dataset 3). D, All video segment predictions of the video-EEG dataset.

Dataset 3 was sourced from in-hospital gold-standard video-EEG monitoring camera systems. In 21 infants without seizures, false detections occurred in 3.4% (365/10,860) of all video segments, with a mean FAR per patient of 3.8% (95% CI: 1.0%-6.5%; Figure 4). As an additional control we assessed videos from one child with ES (45 seizure and 330 non-seizure segments), resulting in an AUC of 0.98, sensitivity of 80%, specificity of 99%, and accuracy 97% (Figure 3), closely matching the derivation and external validation smartphone ES datasets.

Finally, we investigated potential contributing factors to false detections across datasets and within dataset 3. Comparing across datasets, we found that dataset 3, which had the highest FAR, had significantly lower video resolution than the other datasets (p =.006, Xi squared test), with only 2% of video segments having a resolution above 720p, and 21% of videos having low resolution under 480p. Dataset 3 included also night-time footage, which was not present in the other datasets. Excluding night-time videos lowered the total FAR in dataset 3 to 2.8% (283/9937 video segments). Dataset 3 contained long-term, continuous monitoring of children in hospital rooms and was prone to additional sources of bias and obstructions, including EEG caps, bed cribs, and family member interference. To systematically assess the overall visibility of the child within the video, we used an object detection model and compared model confidence measures between videos with and without false detections. This confidence measure acted as a surrogate marker to child visibility. False positive videos showed significantly lower confidence for infant detection (median confidence of 0.23 in correctly detected vs 0.16 in false positive, p < .001, Mann Whitney U test). In examining technical characteristics affecting FAR, false positive videos had significantly lower image sharpness (median Laplacian variance 207 vs. 286 in correctly detected videos, p < .001, Mann-Whitney U test), and significantly lower bitrate (median kilobytes per second of 3496 vs. 3545 in correctly detected videos, p = .006, Mann-Whitney U test). Collectively, these results thus suggest that false alarm rate and seizure detection performance hinge on video quality and child visibility – both of which can be addressed when collecting new data from smartphones or in-hospital video cameras.

## Discussion

Rare disorders are often difficult to diagnose and treat, as they can be paroxysmal in nature and diagnostics tests as well as specialist clinical expertise are often not widely available. Ninety percent of rare disorders in childhood have major neurological effects, and AI technology holds promise in accelerating diagnosis but requires large datasets. Early Identification of seizures is critical for accurate and timely diagnosis of early age onset epilepsy, particularly for IESS, a rare neurological disorder often marked by delayed diagnosis and treatment^5,6,11,12^. With the increasing availability of smartphones and integration of AI into daily life, automated detection of seizures from videos has potential to address this clinical need. Here, we curated a large, heterogeneous dataset of seizure videos from social media to develop and test an AI model capable of detecting ES with high performance. We validated our model across multiple independent datasets, demonstrating strong performance with low FARs. Notably, our model, initially trained on smartphone videos, also showed robust adaptability when applied to gold-standard video-EEG monitoring recordings, though with an increase in FAR in the hospital setting.

AI has been successfully applied to automate multiple tasks in the medical domain, including analysis of diagnostic imaging and electronic health records^21,22^. In epilepsy, AI has accelerated and improved various aspects of care, including EEG analysis, neuroimaging interpretation, seizure detection and forecasting based on data from wearable medical devices, and recently, seizure detection from video-EEG monitoring recordings^23,24^. While advancements in medical AI have been propelled by the development of datasets from various sources such as electronic health records, imaging, and histology, obtaining datasets for rare neurological disorders remains a significant challenge due to the limited availability of data and regulatory constraints on data sharing^21^. Establishing video datasets for medical vision AI is furthermore challenged due to issues related to patient privacy. In epilepsy, open-source datasets of epileptic seizure footage are generally very small and limited to educational purposes. We addressed these challenges by demonstrating the feasibility of collecting a clinically relevant dataset of a rare neurological condition using openly available data from social media. This approach allowed us to identify 167 infants with over 1000 ES. In comparison, a single tertiary center serving a large population of three million people would, on average, see approximately 24 such children per year, thus a similar-sized dataset would be expected to take years to develop^25^. Furthermore, this approach established a diverse, heterogenous cohort, potentially reducing the need for model retraining, improving accessibility across different populations and boosting clinical applicability, addressing a common barrier for AI integration in everyday clinical practice^21^.

Previous studies have begun to explore the use of social media in medical AI development, demonstrating its growing potential. One study used YouTube to create a dataset for analysis of open surgery techniques and assess surgical skills. Another study created an image-based foundational model for pathology based on histological images uploaded to Twitter^26,27^. In neurology, YouTube videos have been used as adjunctive data sources for development or testing of AI models in assessment of Parkinson’s disease, abnormal gait, essential tremor, facial paralysis, and autism(17–22). However, our approach is unique in that it adds to this body of literature by using social media to create a dataset for a rare epileptic disorder, including one of the most sizable datasets in the field of automated video analysis of seizures^23^.

Our ES detection model performed with high accuracy, achieving an AUC of 0.96, sensitivity of 82% and specificity of 90%, as evaluated using a five-fold cross validation approach and similar levels of performance on the out-of-sample test datasets. To develop the AI model, we adapted an existing large scale vision transformer model, which had originally been trained for general action recognition tasks on a massive dataset of videos from the internet^18,19^. Through this we reduced the amount of data required for training, making it a feasible approach for developing a medical AI model with limited training data, such as IESS. Our strategy of adapting a generalist foundational model to medical AI aligns with emerging trends in the field^28^. To our knowledge and based on recent reviews of video-based seizure detection, ours is the first approach utilizing the vision transformer architecture in the domain of video-based seizure detection. Previous methods predominantly relied on skeleton landmark identification, frame-based approaches combined with neural networks, or optical flow methods^23,29^. The vision transformer architecture offers several advantages over these more traditional approaches, including superior ability to capture long-range dependencies in video sequences, enhanced robustness to variations in camera angle and lighting conditions, and improved performance on subtle motion patterns characteristic of ES^30,31^.

According to best practice in the field, we validated our model on three additional out-of-sample datasets. In two independent smartphone derived cohorts, our model performed similarly to the derivation cohort, achieving an AUC of 0.98 on a cohort of infants with ES. Notably, testing on a dataset of normally behaving infants, we found a FAR of 0.75%, with 62 of 67 children having no false detections. This is a key finding with regard to potential clinical applicability, as false alarms are a barrier to clinical adaptation of new diagnostic technology. To note, dataset 2 contained mostly videos from TikTok, a different social media platform from the derivation dataset introducing an additional source of variability that did not affect the model performance.

While our model was trained exclusively on smartphone data, we also tested it on a cohort of infants undergoing gold standard video-EEG (dataset 3), to assess robustness and transferability to other camera sources. The ability of an AI model to perform well on different video sources is a measure of generalizability and could enable translation to different clinical settings. The hospital-based dataset differed from the derivation dataset considerably, as these were long-term videos captured from wide-view stationary cameras that were distant from the subject compared to smartphone videos. Infants wore EEG caps, resolution was significantly lower, there were often additional people (family members, medical staff) in the frame, and videos were recorded both during day and nighttime. Nevertheless, our model achieved a comparable sensitivity on 45 seizures from a patient with ES, and on an additional 21 patients - comprising over 10,860 video segments without seizures – the total FAR was 3.4%. Although this FAR rate is higher than that observed in the smartphone data, it shows promise for potential future utilization in this clinical setting. Testing in future prospective studies should control for these confounders, and future work should focus on incorporating video data from multiple camera sources within seizure detection training data to further improve transferability to different sources of data.

The field of video analysis for seizures in epilepsy is a developing one, with a recent review having identified 34 studies assessing video-based seizure detection^23^. This review revealed large heterogeneity in methods used, dataset size, and performance measures reported. The largest study to date reported the use of a dedicated audio/visual system for seizure detection and included 104 patients and a total of 2767 seizures. This study included multiple seizure types and detailed the clinical decision-making outcome of the intervention, but did not report machine learning performance measures such as sensitivity and specificity, somewhat limiting interpretability^32^. A prior phase 3 study by the same group did report these metrics, however, their approach was semi-automated with involvement of a clinical neurophysiology specialist in the loop^33^. In early onset epilepsy, a notable phase 2 study aimed at detecting clonic seizures examined 12 infants in the hospital setting, and achieved a maximal AUC 0.79 in discriminating a total of 78 clonic seizures from random movements or other seizure types. Other studies focusing on pediatric seizure have shown promise for multiple seizure types including tonic-clonic seizures^24^, nocturnal motor seizures^34^, and neonatal seizures^35–39^. We contribute to this literature by reporting a model trained on smartphone data rather than from in-hospital cameras or specialized hardware, yielding high performance, and assessing seizure detection in the in-field setting. Our study includes a large number of participants and seizures, focuses on ES-a subtle seizure semiology- and adheres to the reporting and testing standards set forth by the International League Against Epilepsy for validation of AI technology for seizure detection devices^40^.

This study has several limitations. First, in the derivation dataset there was no gold standard electrophysiologic or clinical data to confirm the presence of ES. However, videos were reviewed by two expert neurologists trained to identify ES semiology. Likewise, although unlikely, non-motor seizures could not be excluded from interictal videos due to the lack of EEG. However, these would not be expected to be identified through visual inspection by an expert as well, and external validation of the model was done on gold standard confirmed infants with and without epilepsy. Second, since we did not have demographic or clinical data available for social media videos, it was not possible to fully exclude potential biases related to age, sex, or ethnicity. Furthermore, some participants had more video segments than others, and in order to maximize the dataset we could not balance seizures to interictal segments in a 1:1 ratio. To address this limitation, data collected was highly heterogeneous, a large dataset was examined, and additional validation was conducted on completely independent datasets of normally behaving infants and infants with gold-standard video-EEG. Third, while ES are the predominant seizures in children with IESS, they may have additional seizure types, and expansion of our approach to additional seizures is necessary to have true clinical applicability. Fourth, the study design is limited by an inherent selection bias toward videos that were uploaded to social media and future prospective studies are needed to further establish generalizability.

## Conclusion

We here report a novel approach for AI model development suitable for rare neurological disorders. We address the clinical need for early detection of ES in infants by developing a high performing video-AI model capable of identifying this subtle seizure semiology from widely available smartphone videos. The model’s robust performance across multiple datasets, including gold-standard video-EEG recordings, underscores its potential for clinical application. While challenges remain, including further prospective validation in different clinical settings, this work lays a foundation for future developments in video-based automated seizure detection.

## Author Contributions

GM, MH, and CM contributed to collecting, analyzing, and interpreting the data, writing the manuscript, and critically revising the manuscript. GM and CM annotated and classified video segments. MH contributed to the collection of data and critically revising the manuscript. ST contributed to analysis and interpretation of the data, and critically revising the manuscript.

## Competing interests

CM is part of patent applications to detect and predict clinical outcomes and to manage, diagnose, and treat neurological conditions, all of which are outside the submitted work, and declares no othe financial or non-financial competing interests. MH reports personal fees from Angelini, Bial, Desitin, Eisai, Jazz Pharma, Neuraxpharm, Nutricia, and UCB within the last 3 years, outside the submitted work, and declares no non-financial competing interests. GM, MH, ST declare no financial or non-financial competing interests.

## Acknowledgements

Nothing to declare.

## Notes

### Funding Statement

This study was partially funded by a grant from the Berlin Institute of Health.

### Author Declarations

IRB of Charite Universitatsmedizin Berlin gave ethical approval for this work (reference number EA2/273/23).

